# Phenotype-integrated reinterpretation of laboratory-reported *ABCA4* gene sequencing results improves molecular diagnostic rate in Black/non-White patients and those with late-onset Stargardt macular dystrophy

**DOI:** 10.1101/2025.08.28.25334377

**Authors:** Dorothy T Wang, Bani Antonio-Aguirre, Annabelle Pan, Maria Ludovica Ruggeri, Setu P Mehta, Christy H Smith, Kelsey S Guthrie, Carolyn Applegate, Jefferson J Doyle, Mandeep S Singh

## Abstract

While diagnostic disparities by race and age of onset are reported in inherited retinal diseases, their impact on Stargardt disease (STGD)—a clinically and genetically heterogeneous macular dystrophy—remains unclear. We analyzed 246 STGD patients at a U.S. referral center (2003-2024) who completed genetic testing, comparing laboratory-reported results (lab-GT) with manual, phenotype-integrated reinterpretation (m-GT) of *ABCA4* sequencing incorporating updated variant databases and genotype-phenotype correlation. Diagnostic yield and variant burden were assessed by race and age of onset. Positive/likely positive (P/LP) lab-GT was identified in 79% (195), with 78% (191) attributable to *ABCA4* (*ABCA4-*positive lab-GT: 57% [141]). M-GT increased *ABCA4-*P/LP yield to 91% (224). Black participants had lower *ABCA4-* positive lab-GT than Whites (55% vs. 73%) and fewer pathogenic variants; on multivariable analysis, Black race (OR 0.34) and later age of onset (OR 0.95/year) independently predicted reduced molecular diagnosis. The disparity by race resolved with P/LP m-GT (89% vs. 90%); by age of onset, yield remained lower in late-onset cases (*ABCA4-*P/LP lab-GT: 86% early-[≤10yrs], 83% intermediate-[11-44yrs], 54% late-onset [≥45yrs], improving to 97%, 94%, and 77% after m-GT). Post-test reinterpretation improves diagnostic yield, particularly for Black and late-onset STGD patients, underscoring the value of ancestry-informed interpretation, historical reanalysis, and genotype-phenotype correlation.

## Introduction

Stargardt disease (STGD), the most common hereditary macular dystrophy, is usually caused by autosomal recessive *ABCA4* variants. Classic findings include progressive central vision loss, foveal atrophy, and yellow-white flecks in the posterior pole.^1^ Although typically beginning in childhood or adolescence, STGD can present across the lifespan; late-onset cases may resemble age-related macular degeneration (AMD) or other retinal conditions.^1–3^

Next-generation sequencing has improved diagnostic yield for inherited retinal diseases (IRDs) to 56-76%.^4,5^ For STGD, reported yields are typically higher (57-94%), although “solved” case definitions vary, with some requiring biallelic pathogenic variants *in trans*.^6–20^ Over 1,000 disease-associated *ABCA4* variants are known, with allelic heterogeneity driving clinical variability in severity and age of onset.^21,22^ Interpretation is further complicated by novel or uncertain variants and deep intronic alleles that may not be detected by standard genetic testing (GT).^23^

Prior IRD studies suggest that later age of onset and Black race are linked to lower diagnostic yield, but these associations remain understudied in STGD.^6,24–28^

To address these gaps, we evaluated GT in a large single-center STGD cohort. Specifically, we aimed to: (1) compare diagnostic rates using laboratory reports vs. phenotype-integrated reinterpretation of *ABCA4* variants; (2) characterize *ABCA4* variants, including novel alleles; (3) evaluate sociodemographic and clinical factors—including age of onset and race/ethnicity—associated with molecular diagnosis.

## Methods

### Ethics and Participant Selection

This retrospective study was approved by the Johns Hopkins IRB (IRB00213488) with waiver of informed consent, and adhered to the Declaration of Helsinki. Data were de-identified and handled in compliance with HIPAA regulations.

We identified patients seen 2003-2024 at the Wilmer Eye Institute Genetic Eye Diseases (GEDi) center with clinical STGD diagnosed by fellowship-trained IRD specialists. Inclusion required GT completion. Patients without documented suspicion of STGD were excluded.

Sex, race, and Hispanic/Latino ethnicity (HLE) were recorded. Race/HLE was dichotomized for subgroup analysis (Black vs. non-Hispanic White) per literature on lower diagnostic yields in Black IRD patients; individuals identifying as both were categorized as Black.^27^

Clinical variables included age of symptom onset, age at presentation, follow-up duration, family history of confirmed or presumed IRD in any generation, consanguinity, and best-corrected visual acuity (BCVA). BCVA was recorded in Snellen, converted to logarithm of the minimum angle of resolution (logMAR) for analysis, and classified as better- or worse-seeing eye (lower vs. higher logMAR; if identical, the same value was assigned to both). Age of symptom onset was analyzed as a continuous variable and categorized using two frameworks: Pas et al. framework (early-onset ≤10, intermediate-onset 11-44, late-onset ≥45 years)^29^ and 20-year intervals (<20, 20-39, 40-59, ≥60 years).

### Genetic testing and results interpretation

We recorded the timing and scope of GT, including the number of tests and genes analyzed. Laboratory-reported GT (lab-GT) results were extracted directly from clinical laboratory reports and defined as molecular confirmation of any causative gene for retinal dystrophy, based solely on the laboratory’s interpretation without additional reinterpretation.

Separately, *ABCA4*-specific results (m-GT) were re-evaluated by a trained grader through manual review of lab-GT results, incorporating variant databases (April 2025) and in silico predictions; genotype-phenotype consistency was assessed using clinical documentation. “Positive” m-GT required ≥2 pathogenic/likely pathogenic (P/LP) *ABCA4* variants *in trans* (biallelic) or of unknown phase, or prior enrollment in a STGD clinical trial requiring molecular confirmation of *ABCA4-*associated disease for eligibility. “Likely positive” m-GT included cases with one P/LP variant plus one variant of uncertain significance (VUS), two novel/VUS variants *in trans*, or one P/LP variant (or two *in cis*) with a STGD-consistent phenotype as previously reported.^30^ Remaining cases were classified as inconclusive, or negative if no P/LP *ABCA4* variants were identified.

VUS pathogenicity was assessed using ClinVar, Leiden Open Variation Database, gnomAD, Varsome, Franklin, SpliceAI, and MutationTaster. When available, functional and in silico data were used to classify variants as deleterious/damaging (VUS-D) or benign (VUS-B). For each participant, we recorded numbers of total, P/LP, and VUS-D *ABCA4* variants.

### Statistical Analysis

De-identified data were analyzed using Stata v.18. Normality was assessed with Shapiro-Wilk test; non-normal variables used Mann-Whitney U (two groups) or Kruskal-Wallis (multiple groups), and categorical variables used chi-square or Fisher’s exact tests (when expected counts <5). Multivariable logistic regression identified predictors of positive diagnostic yield (positive/likely positive lab-GT) using sex, race, HLE, family history, age of onset, baseline BCVA (better-seeing eye), GT year, and panel size as covariates. Reference groups were defined by the most frequent category. Statistical significance was set at α = 0.05.

## Results

### Participant characteristics

Of 246 STGD participants with GT results, 47% were male (**Table 1**). The cohort was 64% White, 23% Black/African American, and 11% Asian. Median age at presentation was 37 years (interquartile range [IQR]: 21-53); median symptom onset was 22 years (IQR 10-40; mean ± standard deviation [SD]: 26±17; range: 2-69). Family history of IRD was reported in 42%, and parental consanguinity in 3%. Median follow-up was 4.5 years (IQR 2.0-10.2).

**Table 1.** Demographics, clinical characteristics, and genetic testing history. Data presented as % (n), median(IQR), or mean(SD). Denominators for each variable may vary due to missing genetic reports. ^a^Totals may exceed 100% due to multiracial identities. ^b^For participants with multiple tests, the most recent and/or largest panel was recorded. ^c^*Positive* m-GT: ≥2 pathogenic variants in *ABCA4* (*in trans* or unknown phase). *Likely positive*: one pathogenic + one VUS, two novel/VUS variants *in trans*, or one pathogenic (or two *in cis*) with typical STGD phenotype. ^d^Excludes 15 participants who underwent genetic testing and were enrolled in clinical trials requiring molecular confirmation, but whose genetic reports were unavailable. Abbreviations: *BCVA* best-corrected visual acuity; *GT* genetic testing; *IQR* interquartile range; *IRD* inherited retinal disease; *Lab-GT* laboratory-reported genetic testing results, where a positive result indicates molecular confirmation of any retinal dystrophy, regardless of the causative gene; *m-GT* manual phenotype-integrated reinterpretation of *ABCA4*-specific results; *P/LP* pathogenic/likely pathogenic variants; *SD* standard deviation; *STGD* Stargardt disease; *VUS* variant of uncertain significance; *VUS-D* VUS with predicted deleterious effects

| Parameter | Total (N=246) |
| --- | --- |
| **Male sex** | 47% (115 of 246) |
| **Race<sup>a</sup>** |  |
| White | 64% (157 of 246) |
| Black or African American | 23% (56 of 246) |
| Asian | 11% (26 of 246) |
| American Indian or Alaska Native | 3% (7 of 246) |
| Native Hawaiian or Other Pacific Islander | <1% (1 of 246) |
| Other | 4% (11 of 246) |
| **Ethnicity: Hispanic or Latino** | 4% (10 of 246) |
| **Family history of STGD or IRD** | 42% (104 of 246) |
| **History of consanguinity** | 3% (8 of 245) |
| **Age at presentation** | 36.6 (IQR 20.6-53.3) |
| **Age of symptom onset** | 22 (IQR 10-40) |
| **Received GT prior to baseline visit?** | 15% (38 of 246) |
| **Age at GT (years)** | 39.5 (IQR 22 – 57) |
| **Duration of symptoms prior to GT (years)** | 8 (IQR 3-21) |
| **Duration of follow-up (years)** | 4.5 (IQR 2.0-10.2) |
| **Baseline BCVA, better-seeing eye** | 0.602 (IQR 0.176-0.903) |
| **Baseline BCVA, worse-seeing eye** | 0.796 (IQR 0.301-1.097) |
| **Most recent BCVA, better-seeing eye** | 0.875 (IQR 0.3-1.1) |
| **Most recent BCVA, worse-seeing eye** | 1.00 (IQR 0.70-1.30) |
| **Number of genetic tests** |  |
| 1 | 91% (223 of 246) |
| 2 | 8% (20 of 246) |
| 3 | 1% (3 of 246) |
| **Scope of genetic testing (number of genes tested<sup>b</sup>)** |  |
| Targeted *ABCA4* testing (*ABCA4* only) | 7% (17 of 229) |
| Small panel (includes *ABCA4* and limited retinal dystrophy genes) | 31% (70 of 229) |
| Broader retinal dystrophy panel (>30 but <400 genes) | 52% (120 of 229) |
| Large panel (≥400 genes) | 7% (16 of 229) |
| Whole exome/genome sequencing (>20,000 genes) | 3% (6 of 229) |
| **Lab-GT results (all IRD genes)** |  |
| Positive | 70% (173 of 246) |
| Likely positive | 9% (22 of 246) |
| Inconclusive | 15% (38 of 246) |
| Negative | 5% (13 of 246) |
| ***ABCA4* lab-GT results (*ABCA4* only)** |  |
| Positive | 69% (169 of 246) |
| Likely positive | 9% (22 of 246) |
| Inconclusive or Negative in *ABCA4* | 22% (55 of 246) |
| **M-GT results<sup>c</sup>** |  |
| 2+ pathogenic variants in trans (biallelic) | 28.5% (70 of 246) |
| 2+ pathogenic variants in unknown phase | 39.8% (98 of 246) |
| Documented positive & enrolled in trial requiring genetic confirmation | 6.1% (15 of 246) |
| Likely positive (1 pathogenic + 1 VUS or 2 VUS/novel variants in trans) | 11.0% (27 of 246) |
| One pathogenic variant with typical STGD phenotype | 5.7% (14 of 246) |
| Inconclusive | 0.8% (2 of 246) |
| Negative in *ABCA4* | 8.1% (20 of 246) |
| **Number of *ABCA4* variants found (n=231<sup>d</sup>)** |  |
| Total *ABCA4* variants | 2.11 (SD 0.83) |
| P/LP *ABCA4* variants | 1.88 (SD 0.81) |
| P/LP or VUS-D *ABCA4* variants | 2.03 (SD 0.82) |

Baseline median BCVA was 0.60 logMAR (∼Snellen 20/80) in the better-seeing eye and 0.80 (∼20/125) in the worse-seeing eye; among better-seeing eyes, 36% had BCVA 20/40 or better and 13% worse than 20/400. At last follow-up, BCVA declined to 0.88 (∼20/150) and 1.00 (∼20/200) in the better- and worse-seeing eyes, respectively, with 23% having 20/40 or better and 26% <20/400.

### Genetic testing characteristics

Median age at GT was 39.5 years (IQR 22-57), with median symptom duration of 8 years prior to GT (IQR 3-21). Most (91%) had a single Clinical Laboratory Improvement Amendments (CLIA)-certified test, while 9% underwent multiple tests; the most comprehensive and/or most recent result was analyzed. Testing methods included IRD panels (30-400 genes; 52%), smaller panels (<30 genes; 31%), targeted *ABCA4* testing (7%), large IRD panels (>400 genes; 7%), and whole exome/genome sequencing (3%) (**Table 1**). Median GT year was 2019 (IQR 2016-2021; range 2005-2025; **Supplemental Figure 1**). Fifteen percent (n=38) completed GT before their initial evaluation and were excluded from baseline BCVA and age analyses.

### Genetic testing outcomes and classifications

Lab-GT classified 70% (n=173) as positive for any IRD gene, 9% (n=22) likely positive, 15% (n=38) inconclusive, and 5% (n=13) negative (**Table 1**). Of 195 positive/likely positive cases, 191 were *ABCA4*, three *PRPH2*, and one *PROM1*.

Manual phenotype-integrated reinterpretation (m-GT) of *ABCA4* data increased positive classifications to 74% (n=183), including 70 (28%) with biallelic P/LP variants *in trans*, 98 (40%) with ≥2 P/LP variants of unknown phase, and 15 (6%) based on prior STGD clinical trial enrollment requiring molecular confirmation of *ABCA4* disease, although GT reports and variant-level data were unavailable (**Figure 1**, **Table 1**). An additional 41 (17%) were likely positive, 2 (<1%) inconclusive, and 20 (8%) negative (see **Methods** for m-GT criteria).

**Figure 1.**
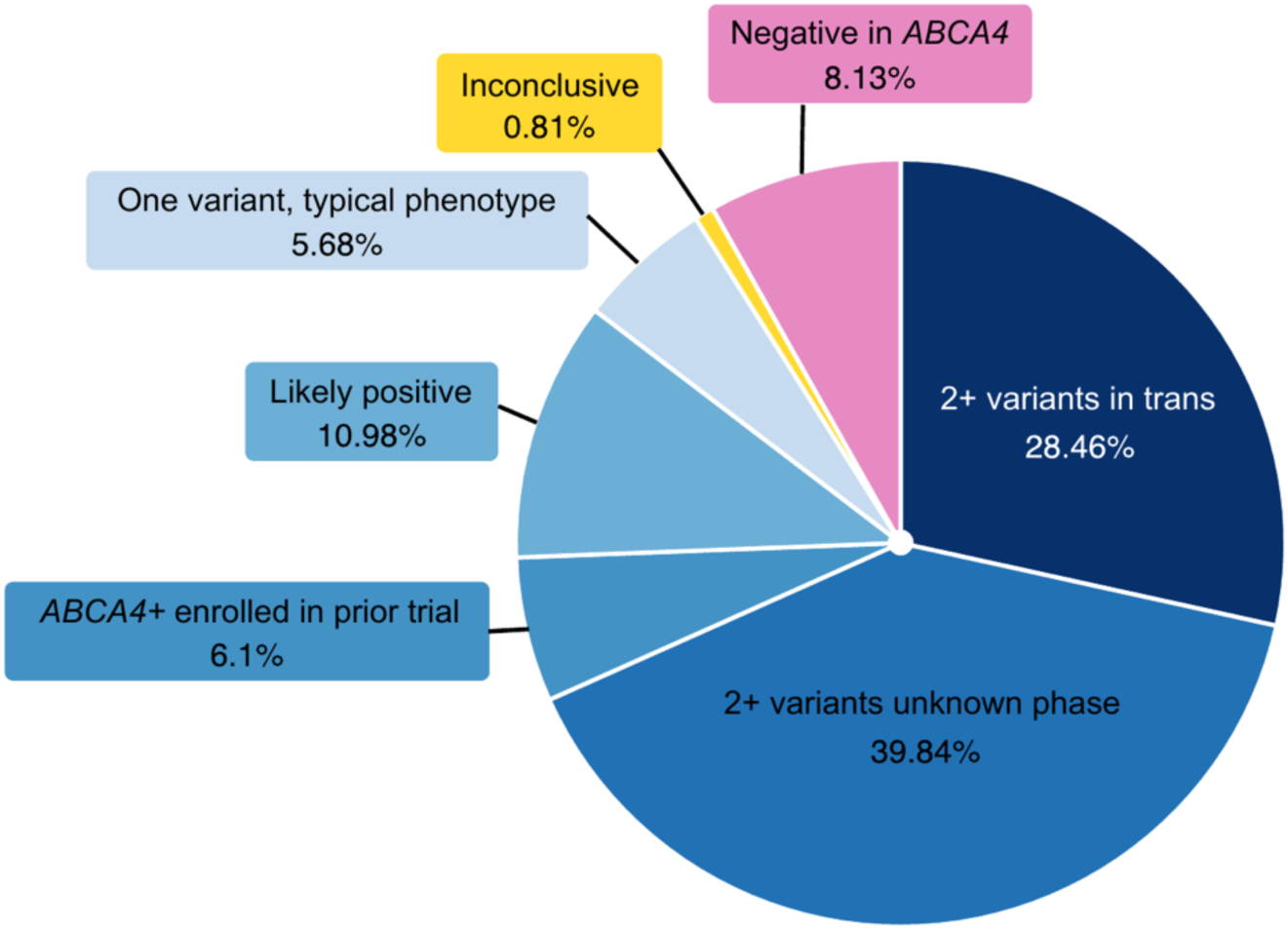
Distribution of per-person *ABCA4* sequencing based on manual reinterpretation of clinical genetic testing results (m-GT; n=246) Manually reinterpreted genetic testing results with respect to *ABCA4* were categorized as follows: **‘***2+ variants in trans*’ (≥2 pathogenic variants confirmed *in trans*); ‘*2+ variants unknown phase*’ (≥2 pathogenic variants with undetermined phase); ‘*ABCA4+ enrolled in prior trial*’ (participants with documented *ABCA4*-related retinopathy via external testing without available reports, all enrolled in prior clinical trials requiring molecular confirmation); ‘*likely positive*’ (one pathogenic variant and one VUS *in trans*, or two VUS/novel variants *in trans*); ‘*one variant, typical phenotype*’ (one pathogenic variant or two variants *in cis* including at least one pathogenic, with a phenotype consistent with Stargardt disease as assessed by a retinal specialist and according to ProgStar criteria^30^); ‘*inconclusive’* (uncertain *ABCA4* contribution, such as one VUS and one risk allele, or two risk alleles); and ‘*negative in ABCA4*’ (no *ABCA4*-related finding or a pathogenic variant identified in a different IRD gene).

Seventeen participants initially classified as inconclusive (n=3) or likely positive (n=14) by lab-GT were upgraded to positive m-GT through manual review, supported by updated variant databases (e.g., VUS or LP to P/LP in ClinVar), published literature, segregation data, genotype-phenotype correlation, and, in select cases, family history (i.e. immediate relative with an identical phenotype, with or without genetic confirmation) (**Supplemental Table 1**). None harbored pathogenic variants in other IRD genes that could explain their phenotype, supporting the revised *ABCA4* diagnosis. The difference in positivity between lab-GT and m-GT reflects these 17 upgraded *ABCA4* cases, four *PRPH2* and *PROM1 lab-GT-*positive cases, and three with positive lab-GT but only likely positive m-GT due to one pathogenic and one VUS *ABCA4* variant without segregation data. Of 38 initially inconclusive lab-GT cases, only 2 (5%) remained inconclusive after m-GT review; 3 (8%) became positive, 26 (68%) likely positive, and 7 (18%) negative, resolving 95% of uncertain cases (**Figure 2**).

**Figure 2.**
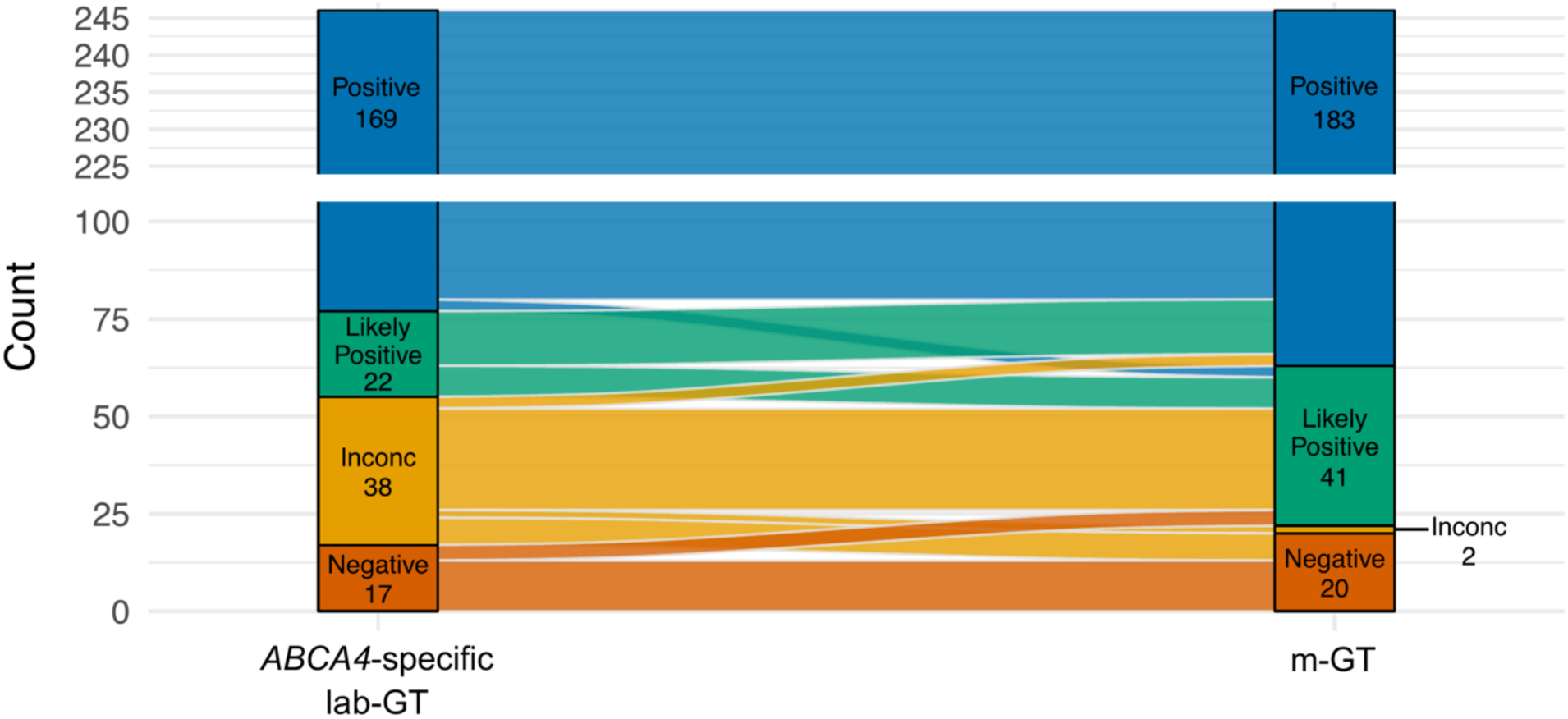
Alluvial plot of *ABCA4-*specific molecular diagnostic classification before and after manual interpretation. Each bar represents the distribution of *ABCA4-*specific genetic testing outcomes before (*ABCA4-*specific lab-GT; genetic testing results in *ABCA4* as directly reported by the laboratory) and after manual gene-specific reinterpretation (m-GT). Colored flows between bars represent participants transitioning between categories. The majority of inconclusive or likely positive lab-GT cases were reclassified to positive or likely positive in *ABCA4* following manual review, while a minority were reclassified *ABCA4-*negative or remained inconclusive (Inconc). Numeric values denote the number of individuals in each category. The “Positive” category is truncated to visually emphasize transitions among likely positive, inconclusive, and negative groups. Note: Four participants had a positive lab-GT result due to pathogenic variants in *PRPH2* (n=3) and *PROM1* (n=1) and were molecularly diagnosed with a non-*ABCA4* inherited retinal disease. For this *ABCA4-*specific analysis, these cases are categorized as “Negative” for *ABCA4* in both lab-GT and m-GT.

### ABCA4 variant spectrum

Among 232 participants with ≥1 *ABCA4* variant (regardless of m-GT classification), 487 variants across 190 unique alleles were identified (**Figure 3, Supplemental Figure 2**): 71% missense, 17% intronic, and 9.5% nonsense/frameshift. Most were P/LP (75%), 14% hypomorphic, and 11% VUS. Hypomorphic alleles, per Lee et al.,^31^ included c.5603A>T (p.N1868I), c.3113C>T (p.A1038V), c.6089>A (p.R2030Q), c.4253+43G>A (p.[=,Ile1377Hisfs*3]), c.4577C>T (p.I1562T), and c.4685T>C (p.W1526M). Hypomorphic/risk allele p.N1868I was most prevalent (allele frequency [AF]: 9.5% [46/487 total *ABCA4* variants]; patient-level frequency [PLF]: 18% [41/232 participants with *ABCA4* variants]), followed by c.5882G>A (p.G1961E; AF 8%; PLF 17%) and c.5461-10T>C (AF 7%; PLF 15%); c.6320G>A (p.R2107H; AF 3.5%; PLF 7%), occurred exclusively in Black participants. (**Figure 3)**.

**Figure 3.**
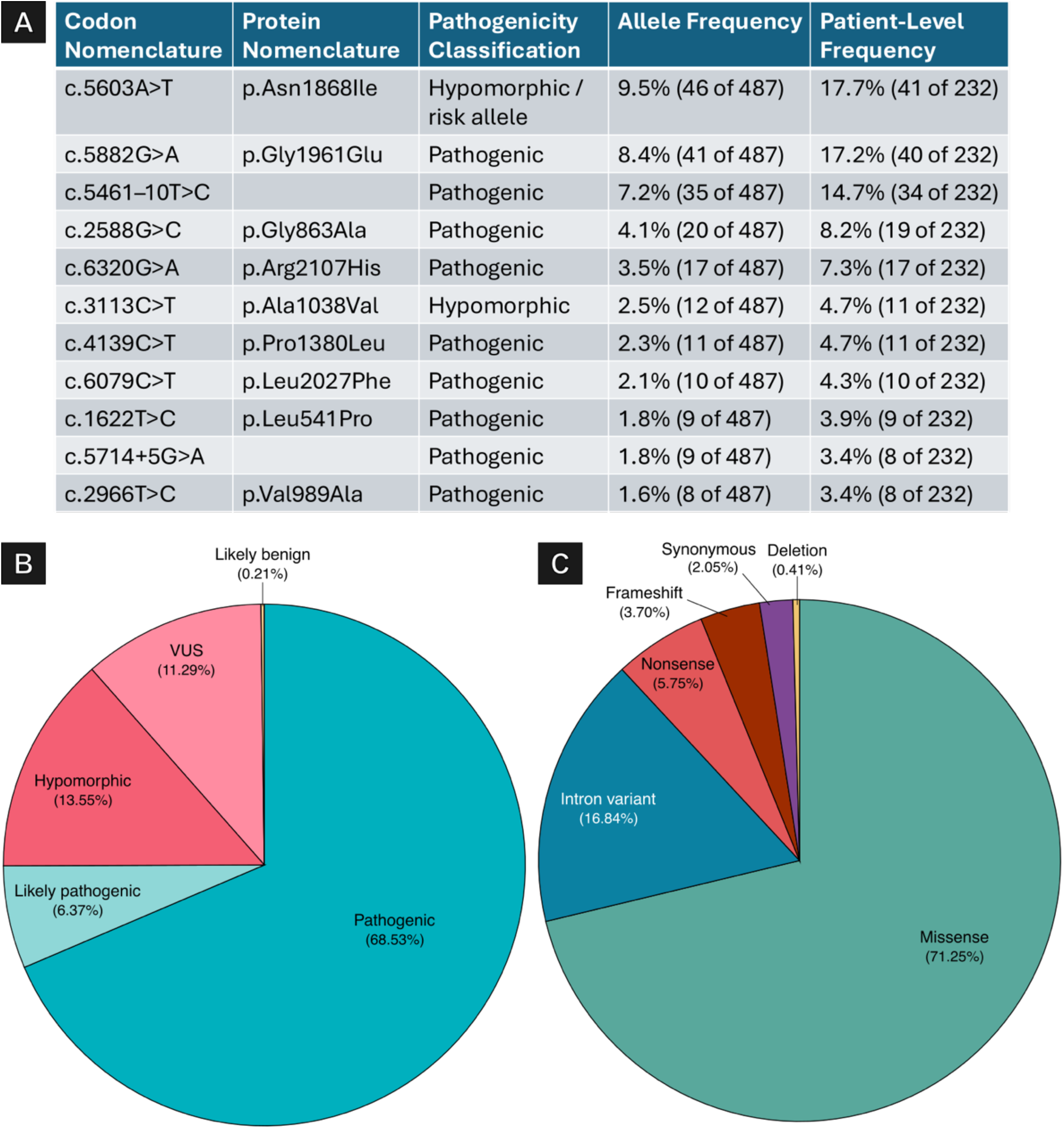
Summary of the 487 ABCA4 variants identified in participants with Stargardt disease. A) 11 most prevalent variants in our cohort. Allele frequency is calculated as the number of occurrences of a given variant divided by the total number of *ABCA4* variants identified; patient-level frequency refers to the number of participants carrying the variant (either heterozygous or homozygous) divided by the total number of participants with *ABCA4* variants. **B)** Distribution of variant-level pathogenicity classifications: pathogenic, likely pathogenic, hypomorphic/risk allele, variant of uncertain significance (VUS), and likely benign. The hypomorphic/risk allele category includes the common allele c.5603A>T (p.Asn1868Ile) and rare hypomorphs such as c.3113C>T, c.4253+43G>A, c.4577C>T, and c.6089G>A. **C)** Distribution of variant consequences, including missense, nonsense, frameshift, deletion, intron variant, and synonymous variants.

Nine variants were novel at the time of GT and remained under-/unreported in public databases as of April 2025 (**Table 2**). All co-occurred with another P/LP allele, though segregation testing was not always performed; most were classified as P/LP, three as VUS. These variants occurred across a broad range of onset ages and diverse racial/ethnic backgrounds, including Black, White, and Asian participants.

**Table 2.** Novel *ABCA4* variants identified in participants with Stargardt disease. Each row represents an *ABCA4* variant classified as novel at the time of genetic testing. Variant details include codon and protein nomenclature, predicted consequence, exon/intron location, zygosity, gnomAD allele frequency, ClinVar clinical significance, and laboratory-reported pathogenicity classification. Zygosity annotations indicate whether variants were confirmed (through parental/segregation testing) or presumed to be *in trans* with another *ABCA4* variant. Ethnicity and age of symptom onset are also reported. All participants harbored at least one additional pathogenic *ABCA4* variant, except for one individual who was homozygous for c.283T>C. Abbreviations: *Het* heterozygous; *Homo* homozygous*; LOVD* Leiden Open Variation Database; *LP* likely pathogenic; *NR* not reported; *P* pathogenic; *VUS* variant of uncertain significance; *VUS-D* VUS with predicted deleterious effects

| Codon Nomenclature | Protein Nomenclature | Variant Consequence | Exon/ Intron | Zygosity | gnomAD Allele Frequency / ClinVar Clinical Significance / LOVD | Laboratory-Reported Pathogenicity Classification |
| --- | --- | --- | --- | --- | --- | --- |
| c.22C>T | p.Gln8* | Nonsense | Exon 1 | Hetero | Not reported / Not found / P | P |
| c.257C>A | p.Thr86Asn | Missense | Exon 3 | Hetero | Not reported / Not found / Not found | LP |
| c.283T>C | p.Ser95Pro | Missense | Exon 3 | Homo | 6.20E-7 / Not found / LP | LP |
| c.1161G>A | p.Trp387* | Nonsense | Exon 9 | Hetero | Not reported / Not found / P | P |
| c.1244A>G | p.Asn415Ser | Missense | Exon 10 | Hetero | Not reported / Pathogenic / LP | VUS-D |
| c.1554G>A | p.Glu518= | Synonymous | Exon 11 | Hetero | 1.24E-6 / Uncertain significance / VUS | VUS-D |
| c.2654-6G>A |  | Intron variant | Intron 17 | Hetero | 6.75E-5 / Conflicting classifications of pathogenicity; Uncertain significance; Likely benign / Not found | VUS |
| c.3931G>T | p.Gly1311* | Nonsense | Exon 27 | Hetero | Not reported / Not found / Not found | P |
| c.6590_6591insGT | p.Gln2198Cysfs*50 | Frameshift | Exon 48 | Hetero | Not reported / Not found / Not found | P |

### Additional genetic findings

Among *ABCA4-*negative participants, 9 (45%) harbored potentially diagnostic variants in other IRD genes (**Supplemental Table 2**). Variants were considered potentially diagnostic if concordant with known disease mechanisms and clinical phenotype (e.g. single P/LP or VUS in autosomal dominant [AD] or X-linked recessive [XL] genes; single heterozygous autosomal recessive [AR] variants were insufficient without additional clinical or molecular evidence). The most frequent gene was *PRPH2*; others included *PROM1*, *EYS*, *CACNA1F,* and *CNGB3.* Both *PRPH2* and *PROM1* are well-described phenocopies of *ABCA4-*associated disease, highlighting the value of multigene panels.^32,33^

Among *ABCA4-*positive participants, 9 carried additional variants in other IRD genes (see above criteria; **Supplemental Table 3**). Most were single variants in AD (*PRPH2, RHO, SNRNP200*) or AD/AR genes (*GUCY2D, RP1L1, RAX2*); one had biallelic VUS in AR gene *CDHR1*. Dual molecular diagnoses were confirmed in three cases (*PRPH2* n=2, *RHO* n=1). Remaining additional variants were classified as VUS after variant database review and lacked sufficient evidence for dual diagnosis. These findings emphasize interpreting IRD sequencing within the context of inheritance and clinical phenotype to avoid over-attributing causality to isolated AR variants or VUS without genotype-phenotype concordance.

### Diagnostic yield by age of onset

Diagnostic yield decreased with later symptom onset (**Figure 4**; all p<0.001 unless otherwise specified). By the Pas framework,^29^ lab-GT positivity for all IRD genes was 75% early-onset (47/63), 77% intermediate-onset (101/131), and 48% late-onset (25/52); including likely positive lab-GT increased yields to 86% (n=54/63), 84% (n=110/131), and 60% (n=31/52), respectively. *ABCA4-*only positive/likely positive lab-GT was 86% (54/63), 83% (109/131), and 54% (28/52). Manual reinterpretation (m-GT) further improved *ABCA4* yield: positive m-GT 84% (n=53/63), 82% (n=107/131), and 44% (n=23/52); including likely positive, 97% (n=61/63), 94% (n=123/131), and 77% (n=40/52).

**Figure 4.**
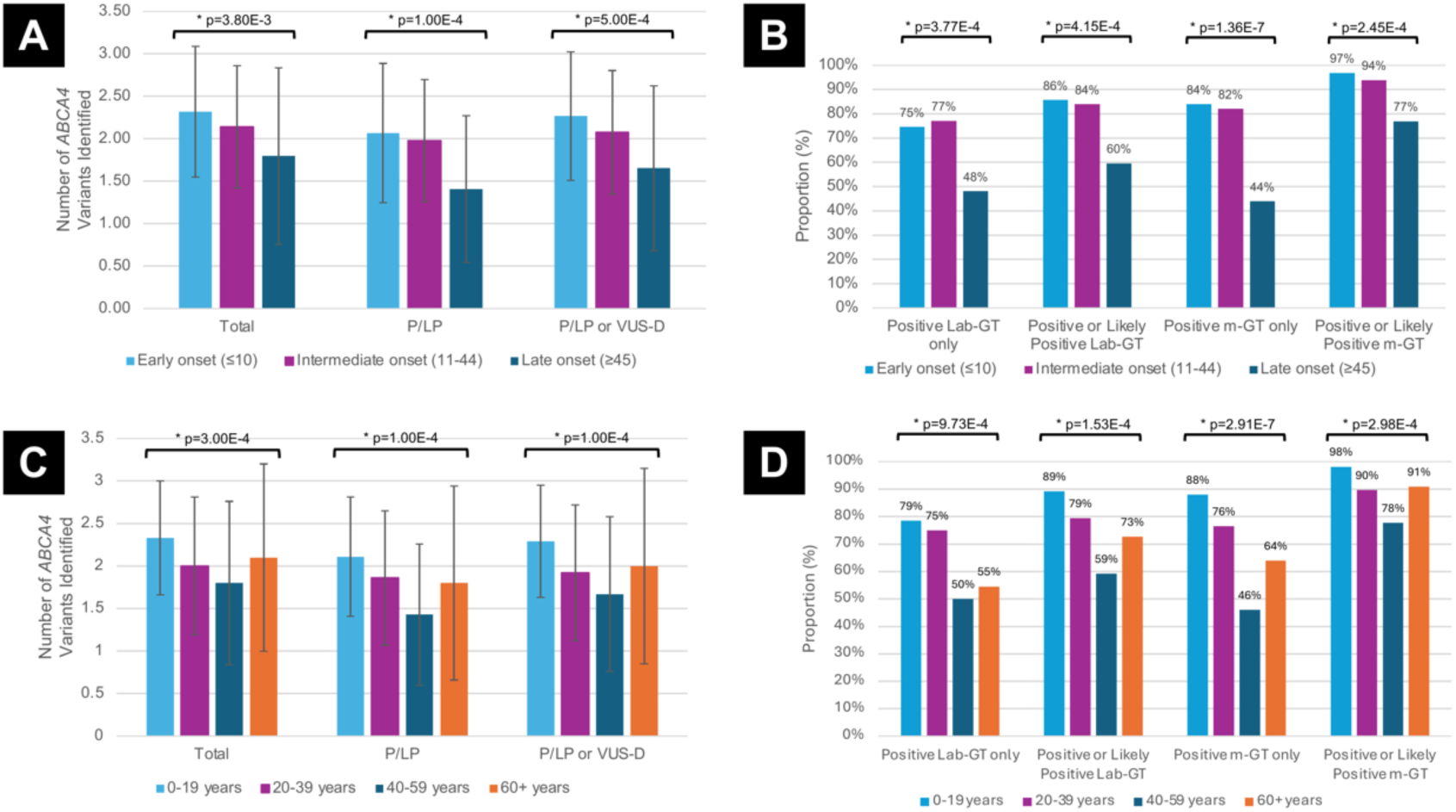
Diagnostic yield by age of symptom onset. A–B) Stratification by the Pas et al. framework (PMID: 37934290): early-onset (≤10 years), intermediate-onset (11-44 years), late-onset (≥45 years). A) Mean (± standard deviation) number of *ABCA4* variants per participant, including total variants, pathogenic/likely pathogenic (P/LP) variants, and combined P/LP + predicted deleterious variants of uncertain significance (VUS-D). Data are shown for 231 participants with available variant data. B) Proportion of participants with positive and likely positive results based on two classification systems: (1) laboratory-reported genetic testing (lab-GT), defined by the clinical laboratory’s original interpretation, and inclusive of any causative gene; and (2) *ABCA4*-specific classification determined by manual review of genetic testing results by a trained grader (m-GT), defined as positive if ≥2 pathogenic *ABCA4* variants (*in trans* or of unknown phase), and likely positive if one pathogenic and one variant of uncertain significance (VUS), two novel/VUS *ABCA4* variants *in trans*, or one pathogenic variant with typical Stargardt disease phenotype. This includes 245 participants, excluding one with missing age of onset. C-D) Same analyses using 20-year age intervals: 0-19, 20-39, 40-59, ≥60 years. Kruskal-Wallis tests were used for comparisons of *ABCA4* variant burden; Fisher’s exact test (20-year framework) and chi-square test (Pas framework) for used for diagnostic yield comparisons.

Twenty-year interval analyses showed a similar pattern: positive lab-GT for all IRD genes was 79% (88/112; <20 years), 75% (51/68; 20-39), 50% (27/54; 40-59), and 55% (6/11; ≥60); including likely positive, 89% (n=100/112), 79% (n=54/68), 59% (n=32/54), and 73% (n=8/11). *ABCA4-*only positive/likely positive lab-GT: 89% (100/112), 78% (53/68), 54% (29/54), and 73% (8/11); positive/likely positive m-GT rates increased to 98% (n=110/112), 90% (n=61/68), 78% (n=42/54), and 91% (n=10/11).

Mean *ABCA4* variant burden of total, P/LP, and P/LP+VUS-D variants declined with later onset in both frameworks. For the Pas framework, mean (±SD) total variants for the early-/intermediate-/late-onset groups were 2.32±0.77, 2.14±0.72, and 1.80±1.04 (p<0.01); for P/LP, 2.07±0.82, 1.98±0.72, and 1.41±0.86; and for P/LP+VUS-D, 2.27±0.76, 2.07±0.73, and 1.65±0.97. For the 20-year bins, mean total variants were 2.33±0.67 (<20 years), 2.00±0.81 (20-39), 1.80±0.96 (40-59), 2.10±1.10 (≥60); P/LP 2.11±0.70, 1.86±0.79, 1.43±0.83, 1.80±1.14; P/LP+VUS-D 2.29±0.66, 1.92±0.80, 1.67±0.91, 2.00±1.15. Baseline BCVA (better-seeing eye) worsened with earlier onset: median logMAR 0.88 (early-), 0.49 (intermediate-), 0.14 (late-onset); by 20-year bins: 0.88, 0.30, 0.18, and 0.10.

### Diagnostic yield by race/ethnicity

Black participants (n=56) had similar total *ABCA4* variant counts as non-Hispanic White participants (n=150; 1.96±1.01 vs. 2.14±0.82; p=0.06) but fewer P/LP (1.57±0.84 vs. 1.97±0.81; p<0.001) and P/LP+VUS-D variants (1.76±0.89 vs. 2.11±0.84; p<0.01), resulting in lower *ABCA4* yields: *ABCA4+* positive lab-GT 55% (31/56) vs. 73% (109/150; p<0.05); positive/likely positive *ABCA4* lab-GT 63% (35/56) vs. 81% (121/150; p<0.01). For all IRD genes, positive lab-GT was 59% (Black) vs. 74% (White; p<0.05), and positive/likely positive was 66% vs. 82% (p<0.05). Positive m-GT rates were lower in Black participants (55% vs. 79%; p<0.001), but after manual interpretation and including likely positive m-GT, yields were similar (89% vs. 90%; p=0.88) (**Figure 5**).

**Figure 5.**
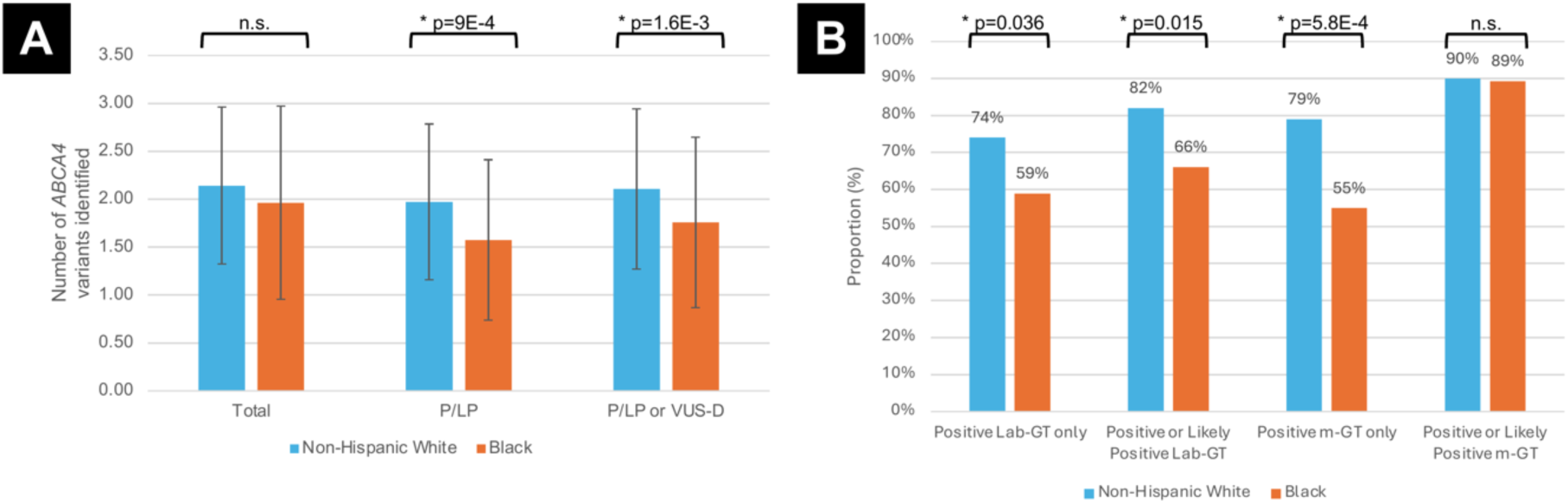
Diagnostic yield by race (Black vs. non-Hispanic White) A) Mean ((± standard deviation) number of *ABCA4* variants per participant, stratified by total variants, pathogenic/likely pathogenic (P/LP) variants, and combined P/LP and predicted deleterious VUS (VUS-D) *ABCA4* variants. This analysis includes 54 Black and 140 non-Hispanic White participants, due to missing variant data. Mann-Whitney tests were used for comparison. **B)** Proportion of participants with positive and likely positive results based on two classification systems: (1) laboratory-reported genetic testing (lab-GT), defined by the clinical laboratory’s original interpretation, and inclusive of any causative gene; and (2) *ABCA4*-specific classification determined by manual review of genetic testing results by a trained grader (m-GT), defined as positive if ≥2 pathogenic *ABCA4* variants (*in trans* or of unknown phase), and likely positive if one pathogenic and one variant of uncertain significance (VUS), two novel/VUS *ABCA4* variants *in trans*, or one pathogenic variant with typical Stargardt disease phenotype. This analysis includes 56 Black participants and 150 White participants. Chi-square tests were used for comparisons.

Baseline BCVA in the better-seeing eye (0.54 logMAR [Black] vs. 0.60 [White]; p=0.79), symptom duration prior to GT (7.5 vs. 7.0 years; p=0.83), and age of onset (30 [IQR 16-44] vs. 21 [IQR 11-42] years; p=0.06) were comparable. Multivariable logistic regression showed Black race (odds ratio [OR]: 0.34; 95% confidence interval [CI]: 0.15-0.79; p<0.05) and later age of onset (OR 0.95 per year; 95% CI 0.92-0.97; p<0.001) independently predicted lower odds of molecular diagnosis (positive/likely positive lab-GT) (**Figure 6, Supplemental Table 4**).

**Figure 6.**
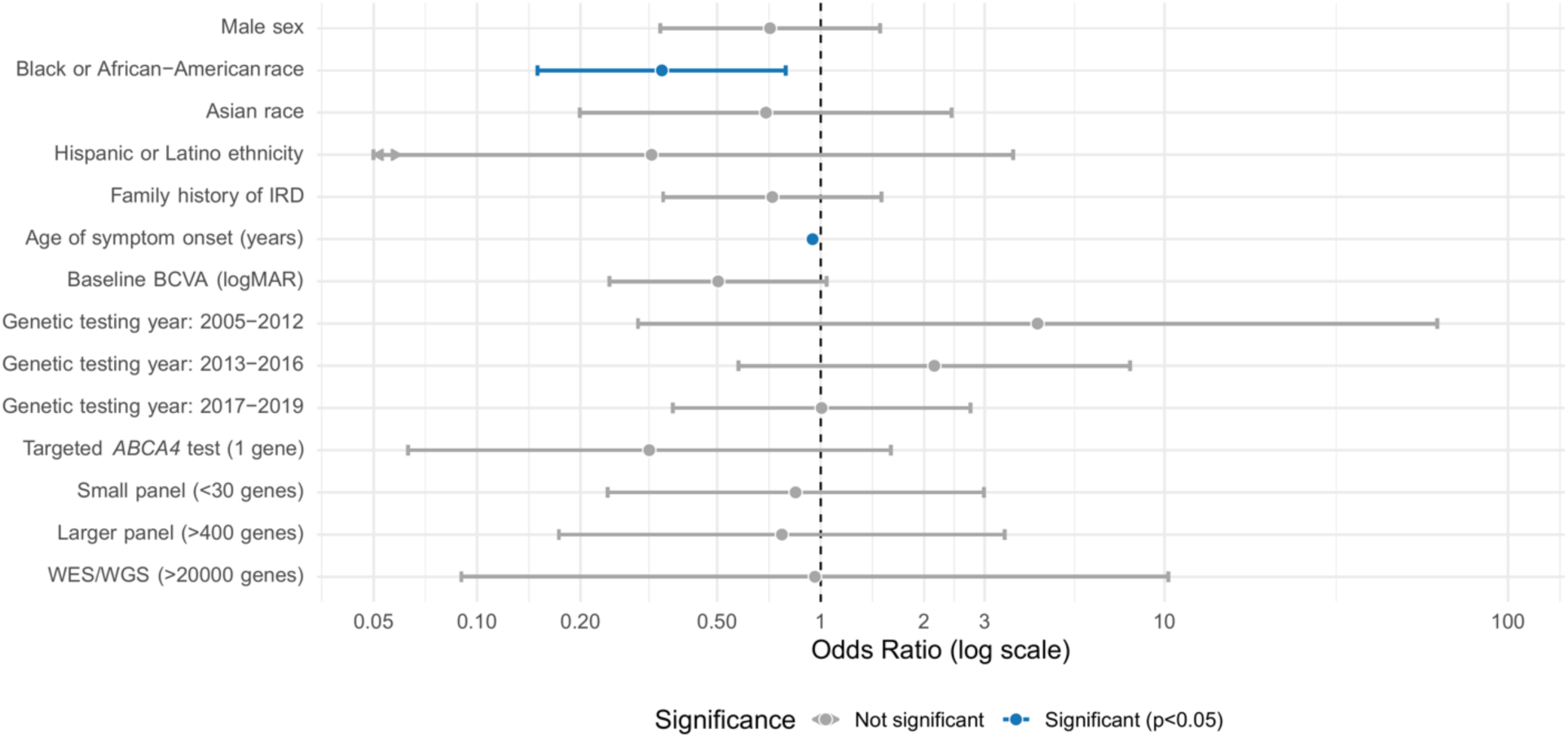
Forest plot showing odds ratios (OR) and 95% confidence intervals (CI) for covariates associated with molecular diagnosis (positive or likely positive lab-GT) in multivariable logistic regression. Black or African-American race and later age of symptom onset were independently associated with significantly lower odds of a molecular diagnosis (p<0.05; shown in blue); all other covariates were not statistically significant (shown in grey). OR are plotted on a logarithmic scale. Arrowheads indicate CI that extend beyond the displayed axis bounds. Reference groups for categorical variables were selected based on the highest frequency group: Female sex, White race, Not Hispanic or Latino, no family history of IRD, genetic testing years 2020-2025, broader retinal dystrophy panel (30-400 genes). See **Supplemental Table 4** for exact OR, CI, and p-values. Abbreviations: *CI* confidence interval; *IRD* inherited retinal disease; *lab-GT* laboratory-reported genetic testing results; *OR* odds ratio

Baseline BCVA, sex, HLE, Asian race, family history, GT year, and panel size were not significant. These findings suggest that diagnostic disparities in Black participants are not fully explained by disease severity or delays in testing.

## Discussion

Our data show that, in Black patients and those with late-onset STGD, manual phenotype-integrated reinterpretation of laboratory-reported *ABCA4* sequencing substantially improved diagnostic yield, raising rates from 78% (positive/likely positive *ABCA4* lab-GT) to 91% (positive/likely positive m-GT) in our large, diverse U.S. cohort. Diagnostic improvements were most pronounced in historically lower-yield subgroups: *ABCA4* yield in Black participants rose from 63% to 90%, matching White participants, and late-onset STGD increased from 54% to 77%.^24,27^ Most previously inconclusive lab results were resolved through VUS reclassification using updated variant evidence, segregation analysis, and genotype-phenotype correlation. While some laboratories routinely update variant interpretations, many do not, underscoring the value of clinician-driven reinterpretation and the limitations of relying solely on lab-issued GT reports for diagnostic purposes in these and potentially other patient groups.

Although Black participants had similar total *ABCA4* variant counts as White participants, they harbored fewer P/LP and VUS-D, leading to lower diagnostic rates by both lab-GT and m-GT. These disparities persisted after adjusting for disease severity and time to GT, reflecting underrepresentation of non-European ancestries in genomic reference databases.^27,34–39^ Together with prior findings that Black IRD patients are less likely to complete GT, these results highlight structural inequities across the diagnostic pipeline and the need for more inclusive reference data and equitable access to GT and counseling services.^40^

The *ABCA4* variant spectrum included globally common alleles (e.g. frequent hypomorph p.N1868I, pathogenic alleles p.G1961E and c.5461-10T>C) and ancestry-specific variants (p.R2107H in Black participants).^41–44^ We also identified nine novel alleles across multiple ancestries despite a majority-White cohort, highlighting the need for ongoing functional validation, population-specific annotation, and routine reanalysis to refine variant classification and diagnostic accuracy.^11,14,17,18,20,45–51,36^

Age of onset predicted both disease severity and diagnostic yield. Early-onset STGD showed higher variant burden, more severe vision loss, and greater likelihood of pathogenic variants, supporting the established association with more penetrant and severe alleles.^24–26,52,53^ Late-onset cases exhibited milder impairment and lower yield, likely reflecting a greater contribution from hypomorphic or deep intronic variants (e.g. p.N1868I or c.4253+43G>A) *in trans* with severe alleles.^31,41^ Many of these variants were historically underrecognized or excluded from earlier panels due to limited intronic region coverage and evolving classification criteria, contributing to underdiagnosis.^23,41,54,55^ A modest rebound in yield was observed among onset ≥60 years (vs. 40-59). Both groups likely include individuals misdiagnosed with non-STGD maculopathies (e.g. pattern dystrophy, AMD with geographic atrophy), due to clinical overlap and lack of routine multimodal imaging.^2,41,54,56,57^ In the ≥60 group, referral to subspecialty IRD care or GT may occur only when the imaging phenotype clearly resembles STGD, creating a selectively referred subset with higher diagnostic yield despite later onset.

The median age of onset was 22 years, slightly older yet still within previously reported ranges (14-22 years), likely reflecting inclusion of more late-onset cases (>20% of our cohort).^44,58–62^ Baseline BCVA supported this phenotypic heterogeneity: 36% had 20/40 or better in the better-seeing eye, while 13% were worse than 20/400, exceeding prior reports of up to 32% and 4%, respectively.^59,61–63^ This spectrum highlights the importance of inclusive research and clinical trial designs that reflect the breadth of STGD presentations.

Our study has several limitations. As a retrospective, single-center study at a U.S. academic medical center, findings may not generalize to other settings. The cohort was predominantly White, potentially limiting applicability to more diverse populations. Inclusion of related individuals may have modestly influenced variant distribution. Changes in GT technology over the study period may have affected diagnostic yield and variant interpretation, particularly in the absence of parental testing or segregation analysis. We also lacked longitudinal visual outcomes and multimodal imaging data collection for detailed phenotypic characterization. Future studies could leverage multimodal imaging and machine learning to explore genotype-phenotype correlations, track *ABCA4* VUS reclassification, and assess whether reanalysis improves diagnostic yield, particularly in other underrepresented populations.

In conclusion, phenotype-integrated reinterpretation of *ABCA4* sequencing markedly enhances STGD diagnostic yield, particularly for Black and late-onset patients. We report one of the highest diagnostic rates to date, identify nine novel *ABCA4* alleles, and demonstrate persistent clinical and molecular heterogeneity across age and racial subgroups. Our findings underscore the diagnostic complexity of *ABCA4* retinopathy and highlight the importance of considering manual re-interpretation of GT results in certain contexts to enhance diagnostic validity.

## Funding Details

This work was supported by the Joseph Albert Hekimian Fund (MSS), Andreas C. Dracopoulos Professorship (MSS), Andreas C. Dracopoulos and Daniel Finkelstein M.D. Rising Professorship in Ophthalmology (JJD), and Research to Prevent Blindness (unrestricted grant to the Wilmer Eye Institute).

## Disclosure of Interest

The authors report there are no competing interests to declare.

## Data Availability Statement

Raw data that support the findings of this study are available from the corresponding author, upon reasonable request.

## Notes

### Competing Interest Statement

The authors have declared no competing interest.

### Author Declarations

IRB of Johns Hopkins University gave ethical approval for this work.

