## Supplemental Figures 1 and 2 for "Phenotype-integrated reinterpretation of laboratory-reported *ABCA4* gene sequencing results improves molecular diagnostic rate in Black/non-White patients and those with late-onset Stargardt macular dystrophy"

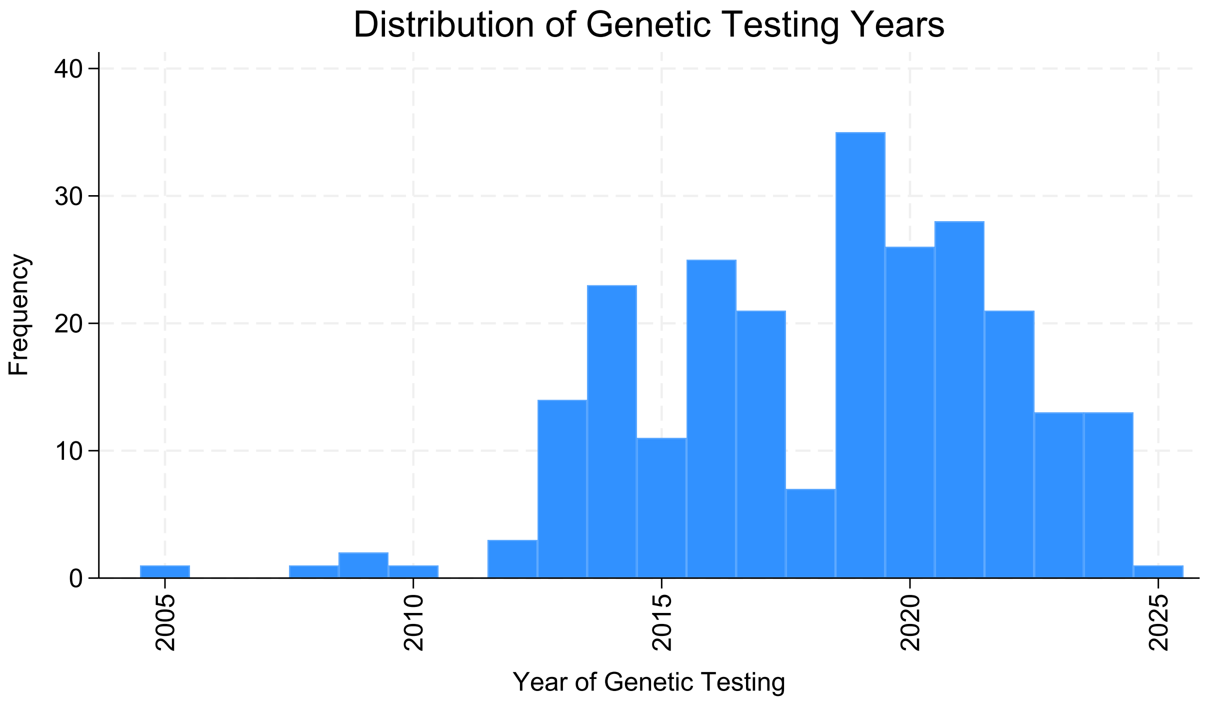


**Supplemental Figure 1. Distribution of genetic testing years among participants with Stargardt disease**

The histogram displays the number of participants who completed genetic testing each year, reflecting temporal trends in testing uptake from 2005 to 2025 within our cohort.

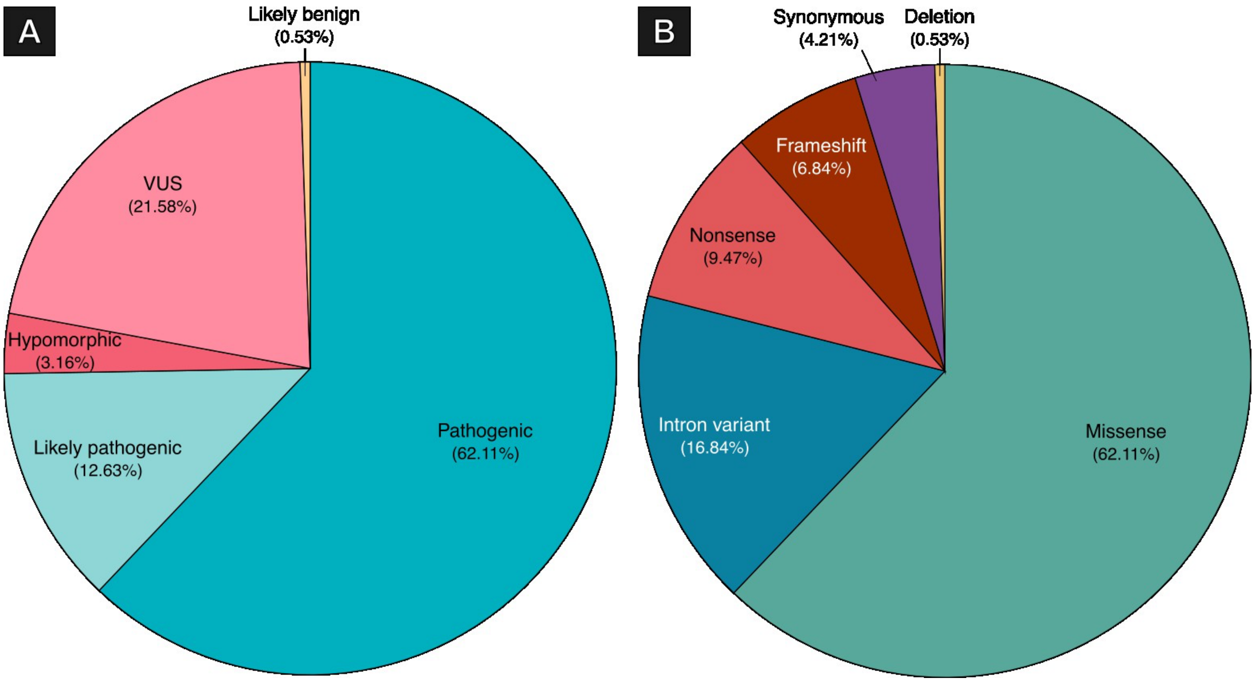


**Supplemental Figure 2. Overview of unique *ABCA4* variants in participants clinically diagnosed with Stargardt disease.**

Of the 487 total *ABCA4* variants identified across our cohort, a total of 190 different variants were found. **A)** Distribution of pathogenicity classifications; **B)** Distribution of mutation types by variant consequence.
