## Supplemental Table 1 for "Phenotype-integrated reinterpretation of laboratory-reported *ABCA4* gene sequencing results improves molecular diagnostic rate in Black/non-White patients and those with late-onset Stargardt macular dystrophy"

Clinical and genetic details for 17 cases upgraded from inconclusive or likely positive lab-GT to positive m-GT. Columns include phenotype, age of onset, genetic testing results, ABCA4 variants, reason for m-GT upgrade (including updated classifications or supporting data), other inherited retinal disease gene findings, and variant-level references.

Abbreviations: *CRD* cone-rod dystrophy; *IRD* inherited retinal disease; *Lab-GT* laboratory-reported genetic testing result (inclusive of all inherited retinal disease genes); *LB* likely benign; *LP* likely pathogenic; *m-GT* manual phenotype-integrated reinterpretation of ABCA4 sequencing results; *MD* macular dystrophy; *P* pathogenic; *PD* pattern dystrophy; *PMID* PubMed ID; *STGD* Stargardt disease; *VUS* variant of uncertain significance

| Case | Pheno-type | Age of Onset (range) | Lab-GT Result | m-GT Result | *ABCA4* Variants at Lab-GT | Reason for m-GT Upgrade | Other IRD Genes? | Supporting References |
|---|---|---|---|---|---|---|---|---|
| P1 | MD/STGD | 20-24 | Inconclusive | Positive: 2+ P/LP unknown phase | 1 P + 1 VUS (c.6320G>A) | VUS largely reclassified as P/LP (ClinVar ID 99448 [Conflicting classifications of pathogenicity; P/LP by 18 submitters]). Varsome VUS-LP; Franklin LP; MutationTaster deleterious. Recognized in the literature as a mild-moderate severity allele associated with African ancestry | None | Lee 2022 (PMID: 34874912), Cornelis 2023 (PMID: 40225145), Zernant 2014 (PMID: 25066811) |
| P2 | STGD | 25-29 | Likely positive | Positive: 2+ P/LP in trans | Novel LP splice site variant + low penetrance P (c.2588G>C; ClinVar ID 7879); in trans on segregation testing | Phase resolved on segregation testing; genotype-phenotype correlation | *TYR* - 1 P, 2 VUS; inconsistent with phenotype | Zernant 2017 (PMID: 28446513), Curtis 2020 (PMID: 32845050), Sangermano 2018 (PMID: 29162642) |
| P3 | CRD/STGD | 5-9 | Inconclusive | Positive: 2+ P/LP unknown phase | 1 P+ 1 VUS (c.4539+2064C>T) | VUS reclassified as P/LP (ClinVar ID 511074). Varsome in-silico predictors benign-supporting; Franklin LP; MutationTaster deleterious; SpliceAI 0.31. Has affected sibling with same *ABCA4* mutations and phenotype (P4) | *TMEM67* (AR gene) - 1 P | |
| P4 | CRD/STGD | 5-9 | Likely positive | Positive: 2+ P/LP unknown phase | 1 P+ 1 VUS (c.4539+2064C>T) | VUS reclassified as P/LP (ClinVar ID 511074). Varsome in-silico predictors benign-supporting; Franklin LP; MutationTaster deleterious; SpliceAI 0.31. Has affected sibling with same *ABCA4* mutations and phenotype (P3) | None | |
| P5 | MD/STGD | 15-19 | Likely positive | Positive: 2+ P/LP in trans | Novel homozygous missense VUS (c.4979C>T) | VUS reclassified as P/LP (ClinVar ID 236123). Varsome VUS-LP; Franklin P; MutationTaster deleterious. | None | |
| P6 | STGD | 15-19 | Likely positive | Positive: 2+ P/LP in trans | 2 LP variants | Both LP classified as P/LP in ClinVar (ID 99266, 99303); phase resolved on segregation testing | None | |
| P7 | STGD | 5-9 | Likely positive | Positive: 2+ P/LP unknown phase | 1 P + 1 VUS (c.4532C>A) | VUS reclassified as P/LP (ClinVar ID 417992). Varsome VUS-LP; Franklin P; MutationTaster deleterious. | None | |

| | | | | | | | | |
|---|---|---|---|---|---|---|---|---|
| P8 | STGD | 5-9 | Likely positive | Positive: 2+ P/LP unknown phase | 1 P + 2 VUS (c.6320G>A, c.468C>T) | Genotype-phenotype correlation; VUS c.6320G>A largely reclassified as P/LP (ClinVar ID 99448 [Conflicting classifications of pathogenicity; P/LP by 18 submitters]); recognized in the literature as a mild-moderate severity allele associated with African ancestry. Other VUS c.468C>T unclear pathogenicity (ClinVar ID 298267; Conflicting classifications of pathogenicity; B/LB by 5 submitters) | None | Lee 2022 (PMID: 34874912), Cornelis 2023 (PMID: 40225145), Zernant 2014 (PMID: 25066811) |
| P9 | STGD/PD | 40-44 | Inconclusive | Positive: 2+ P/LP unknown phase | 1 P + 1 risk allele (c.5603A>T) | Genotype-phenotype correlation (late-onset STGD) | None | Lee 2022 (PMID: 34874912), Zernant 2017 (PMID: 28446513), Garces 2020 (PMID:33375396), Li 2024 (PMID: 37598860) |
| P10 | STGD | 5-9 | Likely positive | Positive: 2+ P/LP unknown phase | 2 LP variants | Both LP classified as P or P/LP in ClinVar (ID 853194, 99428); affected sibling with same phenotype | None | Hu 2020 (PMID: 33129279), Lee 2022 (PMID: 34874912) |
| P11 | CRD/STGD | 5-9 | Likely positive | Positive: 2+ P/LP in trans | 1 P + 1 VUS (c.5311G>A) | VUS reclassified as P/LP (ClinVar ID 1048172). Varsome VUS-LP; Franklin P; MutationTaster deleterious. Phase resolved on segregation testing | None | |
| P12 | STGD | 30-34 | Likely positive | Positive: 2+ P/LP unknown phase | 1 P + low penetrance P (c.2588G>C; ClinVar ID 7879) | Genotype-phenotype correlation | None | Zernant 2017 (PMID: 28446513), Curtis 2020 (PMID: 32845050), Sangermano 2018 (PMID: 29162642) |
| P13 | STGD | 65-69 | Likely positive | Positive: 2+ P/LP unknown phase | 1 P + 1 likely hypomorphic (c.4253+43G>A) | Genotype-phenotype correlation (late-onset STGD); affected sibling with same mutations and phenotype | None | Zernant et al. (2018), Lee 2022 (PMID: 34874912) |
| P14 | STGD | 10-14 | Likely positive | Positive: 2+ P/LP unknown phase | 2 P + 1 VUS (c.5843C>T) | Genotype-phenotype correlation; VUS not considered contributing given presence of 2 other pathogenic variants in *ABCA4* (ClinVar ID 99413; Conflicting classifications of pathogenicity (10 submitters for B/LB)). Varsome VUS-LB; Franklin B; MutationTaster B. | None | |
| P15 | CRD/STGD | 5-9 | Likely positive | Positive: 2+ P/LP unknown phase | 1 P + 1 novel nonsense | Variant reclassified as P/LP (ClinVar ID 861376) | None | |
| A8 | CRD/STGD | 10-14 | Likely positive | Positive: 2+ P/LP in trans | 1 P + 1 novel frameshift | Genotype-phenotype correlation; phase resolved on segregation testing | *RAX2* - 2 VUS | |
| P17 | STGD | 35-39 | Likely positive | Positive: 2+ P/LP unknown phase | 1 P + 1 missense VUS (c.4854G>T) | VUS reclassified as P (ClinVar ID 2910114). Varsome VUS-LP; Franklin P; MutationTaster deleterious. | None | |
