## Supplemental Table 2 for "Phenotype-integrated reinterpretation of laboratory-reported *ABCA4* gene sequencing results improves molecular diagnostic rate in Black/non-White patients and those with late-onset Stargardt macular dystrophy"

Clinical and genetic details for *ABCA4*-negative cases with potentially diagnostic variants in other genes associated with inherited retinal disease (IRD). Columns include phenotype, age of onset, alternative gene(s) and inheritance, detected variants, pathogenicity assessment, clinical comments, and interpretation of causality. Only variants consistent with known patterns and phenotypes were considered potentially diagnostic. Abbreviations: *AD* autosomal dominant; *ar* autosomal recessive; *B* benign; *CACD* central areolar choroidal dystrophy; *ffERG* full-field electroretinography; *iCSNB* incomplete Congenital Stationary Night Blindness; *IRD* inherited retinal disease; *LB* likely benign; *LP* likely pathogenic; *P* pathogenic; *PD* pattern dystrophy; *STGD* Stargardt disease; *VUS* variant of uncertain significance; *XL* X-linked recessive

| Case | Sex | Phenotype | Age of Onset (range) | Alternative IRD Gene (Inheritance) | Variants in alternative genes (as reported on genetic test reports) | Variant Database Information / Pathogenicity for Additional Variants (based on Varsome, Franklin, MutationTaster, ClinVar [when available]) | Clinical / Phenotypic Comments | Causality / Notes |
|---|---|---|---|---|---|---|---|---|
| D1 | Female | STGD | 30-34 | *EYS* (ar) | c.8217delC (LP)<br>c.8220A>T (p.L2740F; VUS)<br>c.6834+59T>C (VUS) | c.8217delC: not in ClinVar; Franklin (LP-P); MutationTaster (deleterious)<br><br>c.8220A>T: not in ClinVar; Varsome/Franklin (VUS); MutationTaster (deleterious)<br><br>c.6834+59T>C: not in ClinVar; Varsome/Franklin (VUS-LB); MutationTaster (B) | Typical STGD phenotype | Per genetic testing report, c.8217delC and c.8220A>T most likely *in cis*; however, no segregation testing has been performed. Possible *EYS* macular dystrophy |
| D2 | Male | STGD | 50-54 | *PRPH2* (AD) | c.457A>G (p.K153E; VUS) | ClinVar 1175280: Conflicting/P/VUS<br>Varsome VUS-LP<br>Franklin: P<br>MutationTaster: deleterious | Typical STGD phenotype | Possible *PRPH2* macular dystrophy |
| D3 | Female | STGD/CACD | 45-49 | *PROM1* (AD/ar) | c.1117C>T (p.R373C; P) | ClinVar ID 5610: P/LP<br>Varsome/Franklin: LP<br>MutationTaster: deleterious | Central areolar choroidal dystrophy with central retinal atrophy | Molecular diagnosis of *PROM1*-related macular dystrophy (Stargardt disease type 4) |
| D4 | Male | STGD | 40-44 | *PRPH2* (AD) | c.515G>A (p.R172Q; P) | ClinVar ID 13167: P/LP<br>Varsome/Franklin: LP<br>MutationTaster: deleterious | Typical STGD phenotype with central geographic atrophy | Molecular diagnosis of *PRPH2*-related dystrophy with central geographic atrophy |
| D5 | Female | STGD | 25-29 | *PRPH2* (AD) | c.136C>T (p.R46X; P) | ClinVar ID 13179: P/LP<br>Varsome: VUS-LP<br>Franklin: P<br>MutationTaster: deleterious | Typical STGD phenotype | Molecular diagnosis of *PRPH2*-related dystrophy |
| D6 | Male | STGD | 5-9 | *CACNA1F* (XL) | c.2982T>G (p.F994L; VUS; NM_005183.4:c.2982T>G) | ClinVar ID 954449: VUS<br>Varsome/Franklin: VUS-LP<br>MutationTaster: B | Fleckless phenotype with central atrophic changes and minimal peripheral pigmentary changes | *CACNA1F* associated with XL iCSNB; unclear if causative/contributory to phenotype, particularly given lack of clear evidence of pathogenicity |

| | | | | | | | | |
|---|---|---|---|---|---|---|---|---|
| D7 | Female | STGD/PD | 50-54 | *PRPH2* (AD) | c.629C>G (p.P210R; P) | ClinVar ID 13173: P/LP<br>Varsome: VUS-LP<br>Franklin: P<br>MutationTaster: deleterious | STGD/PD phenotype with large geographic atrophy | Molecular diagnosis of *PRPH2*-related dystrophy |
| D8 | Male | STGD | 50-54 | *PRPH2* (AD) | c.828+3A>T (P) | ClinVar ID 98713: P<br>Varsome/Franklin: LP<br>MutationTaster: deleterious | Foveal-sparing STGD phenotype | Molecular diagnosis of *PRPH2*-related dystrophy |
| D9 | Female | STGD | 25-29 | *CNGB3* (ar) | c.467C>T (p.S156F; P) | ClinVar ID 427671: Conflicting/P/LP/VUS<br>Varsome/Franklin: LP<br>MutationTaster: deleterious | Presumed STGD, however has symptoms of cone system dysfunction and ffERG suppression of 30Hz flicker amplitude by ~40% | With symptoms of cone system dysfunction and even a single variant in *CNGB3*, may be consistent with *CNGB3*-related achromatopsia |
