## Supplemental Table 3 for "Phenotype-integrated reinterpretation of laboratory-reported *ABCA4* gene sequencing results improves molecular diagnostic rate in Black/non-White patients and those with late-onset Stargardt macular dystrophy"

**Supplemental Table 3.**
Clinical, genetic, and interpretive data for *ABCA4*-positive cases with additional variants in other genes associated with inherited retinal disease (IRD), raising the possibility of dual molecular diagnoses. Columns detail phenotype, age of onset, *ABCA4* genotype, variant(s) found in additional IRD genes, pathogenicity data from laboratory genetic testing reports and current variant databases, clinical comments, and variant interpretation.
Abbreviations: *AD* autosomal dominant; *AF* autofluorescence; *ar* autosomal recessive; *B* benign; *CD* cone dystrophy; *CRD* cone-rod dystrophy; *CSNB* Congenital Stationary Night Blindness; *DDAF* definitely decreased autofluorescence; *ERG* electroretinography; *IRD* inherited retinal disease; *LB* likely benign; *LCA* Leber Congenital Amaurosis; *LP* likely pathogenic; *OMD* occult macular dystrophy; *P* pathogenic; *P/LP* pathogenic/likely pathogenic; *RD* retinal dystrophy; *RP* retinitis pigmentosa; *STGD* Stargardt disease; *VUS* variant of uncertain significance

| Case | Phenotype | Age of Onset (range) | *ABCA4* Genotype (P/LP) | Additional IRD Gene(s) (Inheritance) | Variants in additional genes (as reported on genetic testing report) | Variant Database Information / Pathogenicity for Additional Variants (based on Varsome, Franklin, MutationTaster, ClinVar [when available]) | Clinical / Phenotypic Comments | Causality / Notes |
|---|---|---|---|---|---|---|---|---|
| A1 | STGD/RD | 45-49 | c.2588G>C (p.G863A; P)<br>c.2894A>G (p.N965S; P) | *PRPH2* (AD) | c.866C>T (p.S278L; P) | ClinVar ID 98715: Conflicting/VUS/LB<br>Varsome/Franklin: VUS-LB<br>MutationTaster: deleterious | *ABCA4/PRPH2* fleck retinopathy; mild cone dysfunction | Pathogenic AD variant + *ABCA4*: dual/mixed phenotype |
| A2 | STGD | 10-14 | c.5461-10T>C (homozygous; P) | *SNRNP200* (AD), *CDHR1* (ar) | *SNRNP200*: c.1400T>C (p.L467P; VUS)<br><br>*CDHR1* (in trans): c.1133G>A (P.R378Q; VUS), c.1855G>T (p.V619L; VUS) | *SNRNP200* c.1400T>C: not in ClinVar; Varsome/Franklin (VUS-LP); MutationTaster (deleterious)<br><br>*CDHR1* c.1133G>A: ClinVar 301235 (Conflicting/VUS/LB); Varsome/Franklin (VUS-LB); MutationTaster (B)<br><br>*CDHR1* c.1855G>T: ClinVar 1090074 (Conflicting/VUS/LB); Varsome/Franklin VUS-LB; MutationTaster (B) | Typical STGD phenotype; possible minor *SNRNP200* contribution | Single VUS in AD gene (*SNRNP200*) and biallelic VUS-LB in AR gene (*CDHR1*); insufficient evidence for causality |
| A3 | CRD/STGD | 5-9 | c.4849-1G>A (P)<br>c.6339C>G (p.I2113M; LP) | *GUCY2D* (AD/ar) | c.3181G>A (p.G1061S; VUS) | ClinVar ID 98596: VUS<br>Varsome/Franklin: VUS-LB<br>MutationTaster: B | Severe CRD, some features not classic for *ABCA4* (peripheral bone pigmentation, vessel attenuation) | *GUCY2D* variants typically associated with arLCA, arCSNB, or adCRD; single VUS in AD/ar gene, insufficient to suggest contribution to CRD phenotype given lack of clear pathogenicity |

| | | | | | | | | |
|---|---|---|---|---|---|---|---|---|
| A4 | RP/STGD | 0-4 | c.6320G>A (p.R2107H; P)<br>c.618C>G (p.S206R; LP) | *RHO* (AD) | c.563G>A<br>(p.G188E; P) | ClinVar ID 811432: P/LP<br>Varsome/Franklin: P<br>MutationTaster: deleterious | *ABCA4/RHO*-related retinal dystrophy; macular hyperAF ring and extramacular hypoAF with extensive bone spicule pigment degeneration | Pathogenic AD variant + *ABCA4*: dual/mixed phenotype |
| A5 | STGD | 30-34 | c.5882G>A (p.G1961E; P)<br>c.179C>T (p.A60V; P) | *RP1L1* (AD) | c.1030G>T<br>(p.G344C; VUS) | ClinVar ID 1297657: VUS<br>Varsome/Franklin: VUS<br>MutationTaster: B | Typical STGD phenotype; possible minor *RP1L1* contribution | *RP1L1* associated with AD OMD (and arRP); single VUS in *RP1L1* is insufficient evidence for contribution |
| A6 | STGD/CD | 30-34 | c.6320G>A (p.R2107H; P)<br>c.6079C>T (p.L2027F; P) | *PRPH2* (AD) | c.659G>A<br>(p.R220Q; P) | ClinVar ID 98699: P/LP<br>Varsome/Franklin: P<br>MutationTaster: deleterious | Fleckless bulls'-eye maculopathy (DDAF) - *ABCA4/PRPH2*-related retinal dystrophy | Pathogenic AD variant + *ABCA4*: dual/mixed phenotype |
| A7 | STGD | 10-14 | c.2966T>C (p.V989A; P)<br>c.4634+1del (LP) | *PRPH2* (AD) | c.687C>G<br>(p.N229K; VUS) | Not in ClinVar<br>Varsome/Franklin: VUS-LP<br>MutationTaster: deleterious | Typical STGD phenotype; possible *PRPH2* contribution | Single VUS in AD gene; insufficient evidence for causality |
| A8 | CRD/STGD | 10-14 | c.6590_6591insGT (P)<br>c.1906C>T (p.Gln636Ter; P) | *RAX2* (AD) | *RAX2* (in trans): c.-4A>C (VUS), c.134C>T (p.A45V; VUS; inherited maternally from unaffected mother) | *RAX2* c.134C>T: ClinVar ID 892559 (VUS); Varsome/Franklin (VUS); MutationTaster (deleterious)<br><br>*RAX2* c.-4A>C: not in ClinVar; Varsome/Franklin (VUS-LB); MutationTaster (benign) | Severe CRD with bone spicules, progressive atrophy | *RAX2* variants typically associated with AD CRD; biallelic VUS in *RAX2*; possible contribution but insufficient evidence for causality |
| A9 | STGD | 15-19 | c.1A>G (p.Met1?; P)<br>c.6089G>A (p.R2030Q; P)<br>c.6449G>A (p.Cys2150Y; P) | *GUCY2D* (AD/ar) | c.991C>A<br>(p.R331S; VUS) | ClinVar ID 859779: VUS<br>Varsome/Franklin: VUS-LB<br>MutationTaster: B | Extensive atrophy, pigment changes, attenuated vessels, pending ERG characterization | *GUCY2D* variants typically associated with arLCA, arCSNB, or adCRD; single VUS in AD/ar gene, insufficient to suggest contribution to phenotype given lack of clear pathogenicity |
