## Supplemental Table 4 for "Phenotype-integrated reinterpretation of laboratory-reported *ABCA4* gene sequencing results improves molecular diagnostic rate in Black/non-White patients and those with late-onset Stargardt macular dystrophy"

**Supplemental Table 4.**
Multivariable logistic regression for factors associated with molecular diagnosis. Molecular diagnosis defined as positive or likely positive lab-GT. Odds ratios (OR), 95% confidence intervals (CI), and p-values are shown for all covariates tested, including sex, race, ethnicity, age of symptom onset, family history of confirmed or suspected inherited retinal disease (IRD), baseline best-corrected visual acuity (BCVA) in the better-seeing eye, genetic testing year, and genetic testing panel size. Statistically significant associations ($p<0.05$) are highlighted in bold. The final model included 217 observations after excluding cases with missing data. Multicollinearity was minimal (mean variance inflation factor [VIF]: 1.28), and discrimination was good (area under the Receiver Operating Characteristic [ROC] curve: 0.745).

| Parameter | Odds Ratio (OR) | 95% Confidence Interval (CI) | P-value |
|---|---|---|---|
| **Sex** | | | |
| Male | 0.712 | 0.341-1.489 | 0.367 |
| Female | -- | -- | **--** |
| **Race** | | | |
| White | REF | -- | -- |
| Black or African American | 0.345 | 0.150-791 | **0.012** |
| Native Hawaiian or Other Pacific Islander | -- | -- | -- |
| Asian | 0.693 | 0.199-2.405 | 0.563 |
| Other | -- | -- | **--** |
| **Hispanic or Latino ethnicity** | | | |
| Hispanic or Latino | 0.322 | 0.028-3.633 | 0.359 |
| Not Hispanic or Latino | REF | -- | -- |
| **Family history of confirmed or suspected IRD** | 0.723 | 0.348-1.504 | 0.385 |
| Yes | 0.723 | 0.348-1.504 | 0.385 |
| No | REF | -- | -- |
| **Age of symptom onset (years)** | 0.947 | 0.924-0.971 | **2.00E-05** |
| **Baseline BCVA better-seeing eye (logMAR)** | 0.503 | 0.243-1.040 | 0.064 |
| **Year of genetic testing** | | | |
| 2005-2012 | 4.276 | 0.294-62.239 | 0.288 |
| 2013-2016 | 2.141 | 0.576-7.950 | 0.256 |
| 2017-2019 | 1.006 | 0.371-2.729 | 0.991 |
| 2020-2025 | REF | -- | -- |
| **Genetic testing panel size** | | | |
| Targeted *ABCA4* testing (1 gene) | 0.317 | 0.063-1.599 | 0.164 |
| Small panel including *ABCA4* (<30 genes) | 0.845 | 0.240-2.982 | 0.794 |
| Broader retinal dystrophy panel (30-400 genes) | REF | -- | -- |
| Larger panel (>400 genes) | 0.771 | 0.173-3.426 | 0.732 |
| Whole exome/genome sequencing (>20000 genes) | 0.962 | 0.090-10.258 | 0.975 |
